# Serology in Children with Multisystem Inflammatory Syndrome (MIS-C) associated with COVID-19

**DOI:** 10.1101/2020.07.10.20150755

**Authors:** Christina A. Rostad, Ann Chahroudi, Grace Mantus, Stacey A. Lapp, Mehgan Teherani, Lisa Macoy, Bradley S. Rostad, Sarah S. Milla, Keiko M. Tarquinio, Rajit K. Basu, Carol Kao, W. Matthew Linam, Matthew G. Zimmerman, Pei-Yong Shi, Vineet D. Menachery, Matthew E. Oster, Sri Edupuganti, Evan J. Anderson, Mehul Suthar, Jens Wrammert, Preeti Jaggi

## Abstract

**Objectives:** We aimed to measure SARS-CoV-2 serologic responses in children hospitalized with multisystem inflammatory syndrome (MIS-C) compared to COVID-19, Kawasaki Disease (KD) and other hospitalized pediatric controls.

**Methods:** From March 17, 2020 - May 26, 2020, we prospectively identified hospitalized children at Children’s Healthcare of Atlanta with MIS-C (n=10), symptomatic PCR-confirmed COVID-19 (n=10), KD (n=5), and hospitalized controls (n=4). With IRB approval, we obtained prospective and residual blood samples from these children and measured SARS-CoV-2 spike (S) receptor binding domain (RBD) IgM and IgG binding antibodies by quantitative ELISA and SARS-CoV-2 neutralizing antibodies by live-virus focus reduction neutralization assay. We statistically compared the log-transformed antibody titers among groups and performed correlation analyses using linear regression.

**Results:** All children with MIS-C had high titers of SARS-CoV-2 RBD IgG antibodies, which correlated strongly with neutralizing antibodies (R^2^=0.667, P<0.001). Children with MIS-C had significantly higher SARS-CoV-2 RBD IgG antibody titers (geometric mean titer [GMT] 6800, 95%CI 3495-13231) than children with COVID-19 (GMT 626, 95%CI 251-1563, P<0.001), children with KD (GMT 124, 95%CI 91-170, P<0.001) and other hospitalized pediatric controls (GMT 85 [all below assay limit of detection], P<0.001). All children with MIS-C also had detectable RBD IgM antibodies, indicating recent SARS-CoV-2 infection. RBD IgG titers correlated with erythrocyte sedimentation rate (ESR) (R^2^=0.512, P<0.046) and with hospital and ICU lengths of stay (R^2^=0.590, P=0.010).

**Conclusion:** Quantitative SARS-CoV-2 RBD antibody titers may have a role in establishing the diagnosis of MIS-C, distinguishing it from other similar clinical entities, and stratifying risk for adverse outcomes.

**Table of Contents Summary:** Children with MIS-C have high antibody titers to the SARS-CoV-2 spike protein receptor binding domain, which correlate with neutralization, systemic inflammation, and clinical outcomes.

**What’s Known on This Subject:** Although the clinical features of a multisystem inflammatory syndrome in children (MIS-C) associated with COVID-19 have been recently described, the serologic features of MIS-C are unknown.

**What This Study Adds:** In this case series, all hospitalized children with MIS-C had significantly higher SARS-CoV-2 binding and neutralizing antibodies than children with COVID-19 or Kawasaki Disease. SARS-CoV-2 antibodies correlated with metrics of systemic inflammation and clinical outcomes, suggesting diagnostic and prognostic value.

## Introduction

The majority of SARS-CoV-2 infections in children are either mildly or asymptomatic, with a small subset requiring hospitalization.^1^ However, clusters of a severe inflammatory syndrome in children have recently been reported following the onset of SARS-CoV-2 transmission in areas with a high prevalence of COVID-19.^2-7^ Many of these children have had clinical features similar to Kawasaki Disease (KD)^6-8^ and they have had high rates of hemodynamic instability, myocardial involvement,^9^ and respiratory failure requiring intensive care level support. On May 14, 2020, the U.S. Centers for Disease Control and Prevention (CDC) released a health advisory and a preliminary case definition for this clinical entity, which they termed Multisystem Inflammatory Syndrome in Children (MIS-C) associated with COVID- 19._10_ Although the clinical features of MIS-C have been described, the SARS-CoV-2 serologic responses in these children and in children hospitalized with COVID-19 are unknown. In this study, we measured antibodies to the SARS-CoV-2 spike (S) protein receptor binding domain (RBD) and neutralizing antibodies in children hospitalized with MIS-C compared to COVID-19, KD, and other hospitalized controls.

## METHODS

### Setting and Patient Population

Children and young adults (0 to 21 years of age) hospitalized at Children’s Healthcare of Atlanta from March 17, 2020 to May 26, 2020 were eligible for enrollment for prospective and/or residual blood collection. With IRB approval, samples were collected prospectively when parental consent was possible and residual samples were obtained if consent was not possible. Demographic, clinical, laboratory, and outcome data were obtained through abstraction of the electronic medical record and recorded on a standardized case report form.

### Definition of Cohorts

#### MIS-C

Children who met the CDC case definition were classified as having MIS-C if they had SARS-CoV-2 nucleocapsid (N) protein IgG antibodies detected by a clinician-ordered, qualitative, commercially available assay (Abbott) or if they had nasopharyngeal (NP) detection of SARS CoV-2 by real-time PCR (RT-PCR).

#### COVID-19

Symptomatic hospitalized children who tested positive for SARS-CoV-2 by NP RT-PCR and did not meet the CDC case definition for MIS-C were classified as having COVID-19.

#### KD and other hospitalized controls

Children were classified as having complete or incomplete KD using American Heart Association (AHA) criteria.^11^ Additional contemporaneous hospitalized children who tested negative for SARS-CoV-2 were included as controls.

### SARS-CoV-2 ELISAs

We previously described the cloning, expression, and purification of a recombinant form of the spike (S) RBD from SARS-CoV-2.^12^ We measured anti-RBD antibodies by enzyme-linked immunosorbent assays (ELISAs) in duplicate as described.^12^ Secondary antibodies used for development were anti-hu-IgG-HRP and anti-hu-IgM-HRP (Jackson Immuno Research). Plates were developed using o-phenylenediamine substrate and absorbance was read at 490 nm. Absorbance curves were generated by non-linear regression analysis, and endpoint titers were interpolated from curves using a baseline value calculated from the pooled sera of eight healthy controls. The assay limit of detection (LOD) was 100, and undetectable titers were assigned a value of 85.

### Focus Reduction Neutralization Assays

An infectious clone of the full-length mNeonGreen SARS-CoV-2 (2019- nCoV/USA_WA1/2020) was generated as previously described.^13^ Serially diluted patient serum or plasma and virus were combined and incubated at 37°C for 1 hour. The antibody-virus mixture was aliquoted onto a monolayer of VeroE6 cells and incubated at 37°C for 1 hour. The inoculum was removed and medium supplemented with 1% methylcellulose was added. Plates were incubated at 37°C for 24 hours, and were then fixed with 2% paraformaldehyde. Green fluorescent foci were visualized using an ELISPOT reader (CTL). GFP-based FRNT_50_ curves were generated by non-linear regression analysis. Titers were expressed as the serum dilution at which fluorescent foci were reduced by 50% (FRNT_50_). The assay LOD was defined by the starting dilution of serum samples, which was either 1:20 or 1:40. For graphing purposes, the LOD was represented as 40, and all undetectable titers were assigned a value of 20.

### Statistical Methods

Statistical comparisons were made using GraphPad Prism version 8.0. Frequencies were compared using Chi-squared tests, and continuous variables were compared using one-way analysis of variance (ANOVA) with Tukey’s post-hoc analysis. Linear regression was performed on log-transformed RBD IgG titers or neutralization titers, and the coefficients of determination (R^2^) values were determined. *P*-values ≤ 0.05 were considered statistically significant.

## RESULTS

### Baseline Characteristics

During the study period, we identified 10 children hospitalized with MIS-C, 10 with COVID-19, 5 with KD (Table 1), and 4 hospitalized controls who had serum available for analysis. Children with MIS-C had a median age of 8.5 years (IQR 7-11.5 years), and the majority were male sex, Black race, non-Hispanic ethnicity, and previously healthy with a normal BMI. Distinguishing laboratory features of MIS-C included lymphopenia and thrombocytopenia, and gastrointestinal symptoms were predominant at the time of presentation. Children with MIS-C were significantly more likely to require admission to the intensive care unit (P=0.005) and to have fluid-refractory shock requiring inotropes or vasopressors (P<0.001) compared to children with COVID-19 or KD. All children with MIS-C and KD were ultimately discharged home. However, one child with COVID-19 died from complications of acute myelogenous leukemia and another child remained hospitalized at the time of publication for management of underlying comorbidities.

**Table 1.**
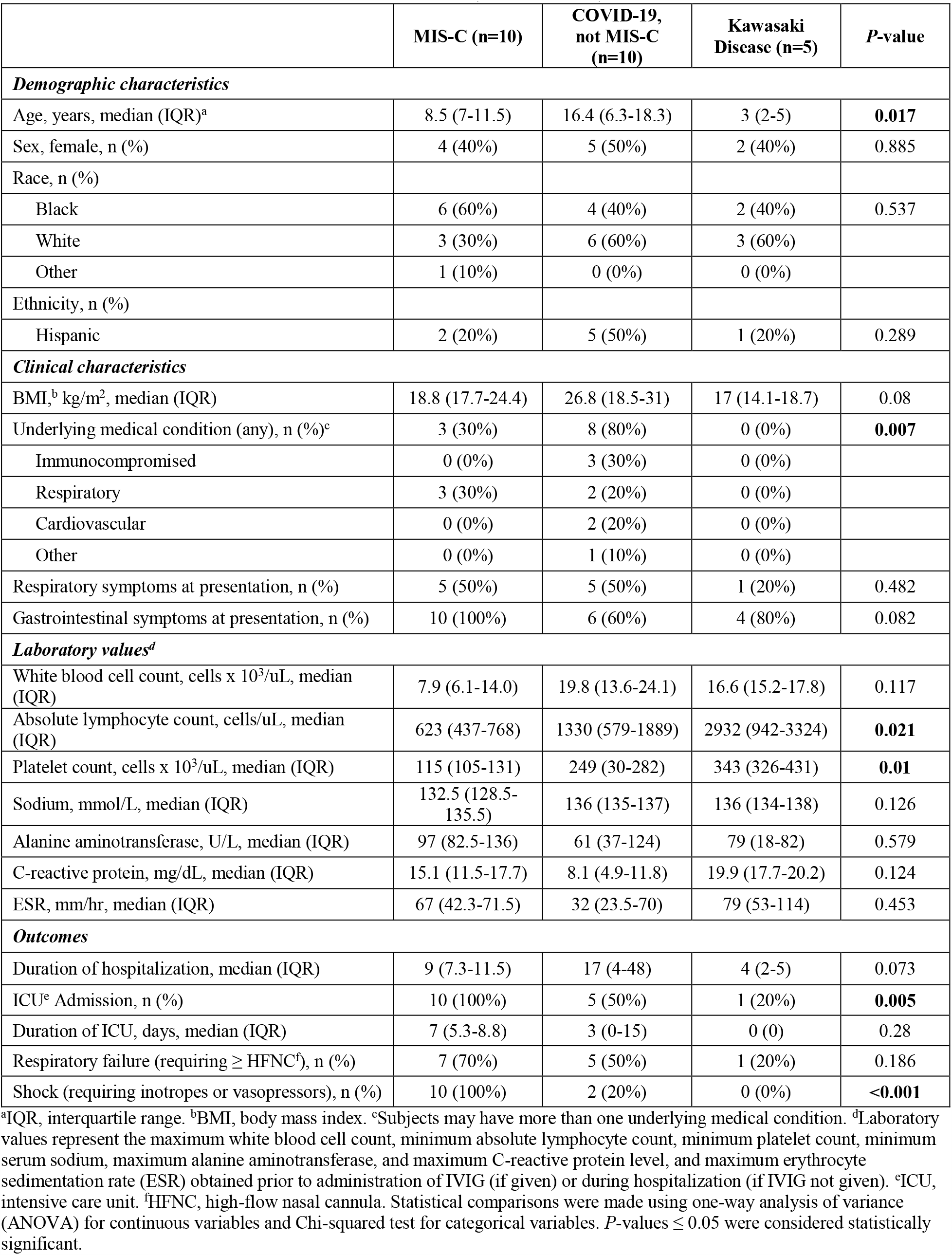
Characteristics of Children with MIS-C, COVID-19, and Kawasaki Disease.

|  | MIS-C (n=10) | COVID-19,<br>not MIS-C<br>(n=10) | Kawasaki<br>Disease (n=5) | P-value |
| --- | --- | --- | --- | --- |
| **Demographic characteristics** |  |  |  |  |
| Age, years, median (IQR)<sup>a</sup> | 8.5 (7-11.5) | 16.4 (6.3-18.3) | 3 (2-5) | **0.017** |
| Sex, female, n (%) | 4 (40%) | 5 (50%) | 2 (40%) | 0.885 |
| Race, n (%) |  |  |  |  |
| Black | 6 (60%) | 4 (40%) | 2 (40%) | 0.537 |
| White | 3 (30%) | 6 (60%) | 3 (60%) |  |
| Other | 1 (10%) | 0 (0%) | 0 (0%) |  |
| Ethnicity, n (%) |  |  |  |  |
| Hispanic | 2 (20%) | 5 (50%) | 1 (20%) | 0.289 |
| **Clinical characteristics** |  |  |  |  |
| BMI,<sup>b</sup> kg/m<sup>2</sup>, median (IQR) | 18.8 (17.7-24.4) | 26.8 (18.5-31) | 17 (14.1-18.7) | 0.08 |
| Underlying medical condition (any), n (%)<sup>c</sup> | 3 (30%) | 8 (80%) | 0 (0%) | **0.007** |
| Immunocompromised | 0 (0%) | 3 (30%) | 0 (0%) |  |
| Respiratory | 3 (30%) | 2 (20%) | 0 (0%) |  |
| Cardiovascular | 0 (0%) | 2 (20%) | 0 (0%) |  |
| Other | 0 (0%) | 1 (10%) | 0 (0%) |  |
| Respiratory symptoms at presentation, n (%) | 5 (50%) | 5 (50%) | 1 (20%) | 0.482 |
| Gastrointestinal symptoms at presentation, n (%) | 10 (100%) | 6 (60%) | 4 (80%) | 0.082 |
| **Laboratory values<sup>d</sup>** |  |  |  |  |
| White blood cell count, cells x 10<sup>3</sup>/uL, median (IQR) | 7.9 (6.1-14.0) | 19.8 (13.6-24.1) | 16.6 (15.2-17.8) | 0.117 |
| Absolute lymphocyte count, cells/uL, median (IQR) | 623 (437-768) | 1330 (579-1889) | 2932 (942-3324) | **0.021** |
| Platelet count, cells x 10<sup>3</sup>/uL, median (IQR) | 115 (105-131) | 249 (30-282) | 343 (326-431) | **0.01** |
| Sodium, mmol/L, median (IQR) | 132.5 (128.5-135.5) | 136 (135-137) | 136 (134-138) | 0.126 |
| Alanine aminotransferase, U/L, median (IQR) | 97 (82.5-136) | 61 (37-124) | 79 (18-82) | 0.579 |
| C-reactive protein, mg/dL, median (IQR) | 15.1 (11.5-17.7) | 8.1 (4.9-11.8) | 19.9 (17.7-20.2) | 0.124 |
| ESR, mm/hr, median (IQR) | 67 (42.3-71.5) | 32 (23.5-70) | 79 (53-114) | 0.453 |
| **Outcomes** |  |  |  |  |
| Duration of hospitalization, median (IQR) | 9 (7.3-11.5) | 17 (4-48) | 4 (2-5) | 0.073 |
| ICU<sup>e</sup> Admission, n (%) | 10 (100%) | 5 (50%) | 1 (20%) | **0.005** |
| Duration of ICU, days, median (IQR) | 7 (5.3-8.8) | 3 (0-15) | 0 (0) | 0.28 |
| Respiratory failure (requiring ≥ HFNC<sup>f</sup>), n (%) | 7 (70%) | 5 (50%) | 1 (20%) | 0.186 |
| Shock (requiring inotropes or vasopressors), n (%) | 10 (100%) | 2 (20%) | 0 (0%) | **<0.001** |

<sup>a</sup>IQR, interquartile range. <sup>b</sup>BMI, body mass index. <sup>c</sup>Subjects may have more than one underlying medical condition. <sup>d</sup>Laboratory values represent the maximum white blood cell count, minimum absolute lymphocyte count, minimum platelet count, minimum serum sodium, maximum alanine aminotransferase, and maximum C-reactive protein level, and maximum erythrocyte sedimentation rate (ESR) obtained prior to administration of IVIG (if given) or during hospitalization (if IVIG not given). <sup>e</sup>ICU, intensive care unit. <sup>f</sup>HFNC, high-flow nasal cannula. Statistical comparisons were made using one-way analysis of variance (ANOVA) for continuous variables and Chi-squared test for categorical variables. P-values ≤ 0.05 were considered statistically significant.

### Serologic Responses

All children with MIS-C and the majority of children with COVID-19 (90%) had detectable IgG antibodies to SARS-CoV-2 S protein RBD (Fig. 1A). The child with COVID-19 who had undetectable antibodies had only mild respiratory symptoms. In contrast, children with KD had minimally reactive antibodies, and all hospitalized controls had undetectable antibody responses. When statistically compared, children with MIS-C had significantly higher SARS-CoV-2 antibody titers (P<0.001) than children with COVID-19, KD, and other hospitalized controls.

**Figure 1.**
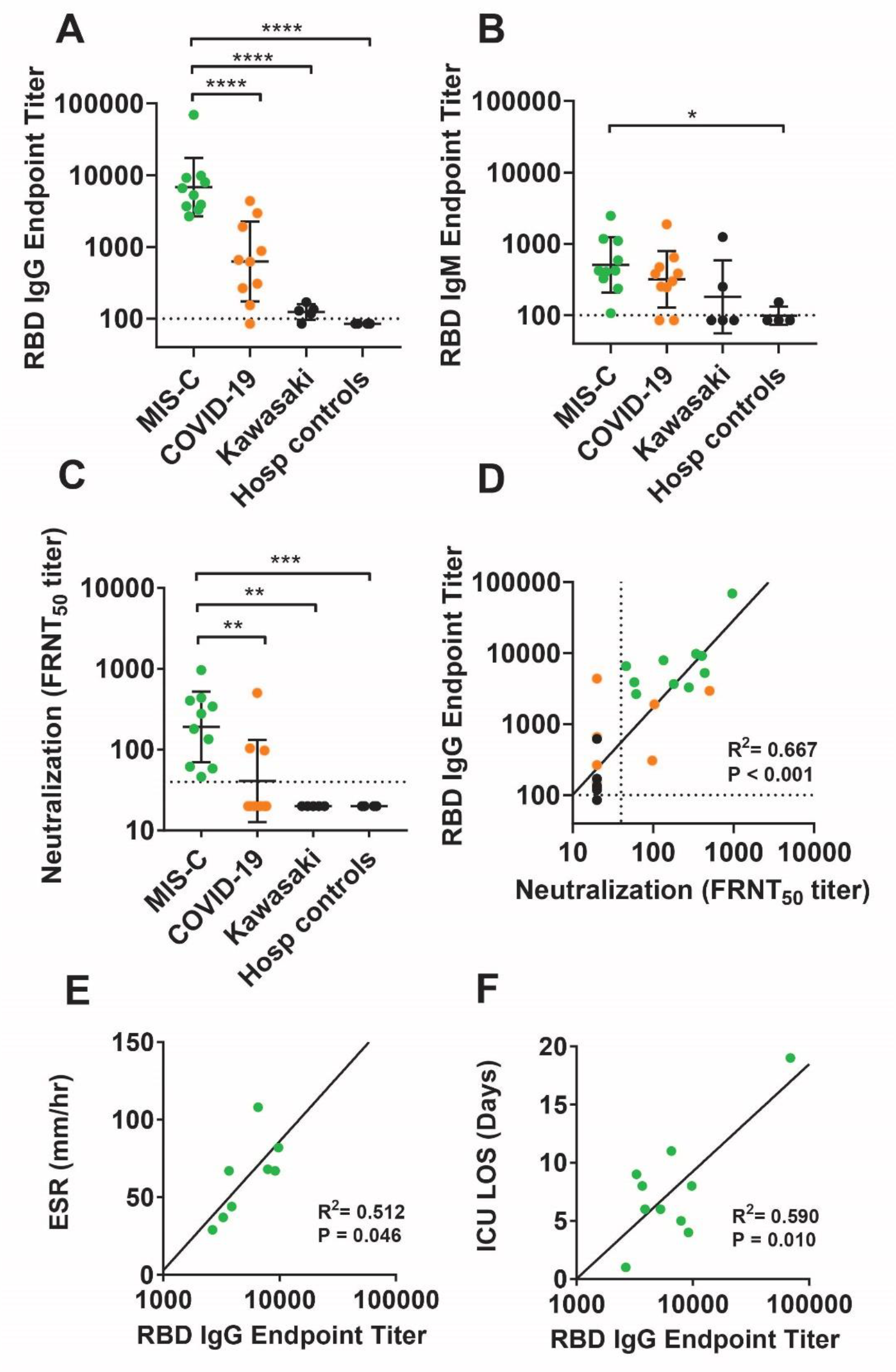
SARS-CoV-2 serology in children with MIS-C. ELISAs to the SARS-CoV-2 RBD, expressed as (A) IgG and (B) IgM end-point titers. (C) SARS-CoV-2 neutralizing antibody titers, expressed as the FRNT_50_. (D) Correlation between log-transformed RBD IgG titers and FRNT_50_; (E) ESR (mm/hr), n=8; and (F) ICU LOS by linear regression.

Similarly, all children with MIS-C and most children with COVID-19 (80%) had detectable IgM antibodies SARS-CoV-2 RBD (Fig. 1B). Although children with MIS-C had significantly higher IgM antibody titers than hospitalized control patients, there were not statistical differences between the other groups. Interestingly, one child with KD had a high RBD IgM titer and negative IgG titer. Although this child had tested negative for SARS-CoV-2 by NP PCR and by nucleoprotein antibody assay (Abbott), her parent did report having a respiratory illness two weeks preceding the child’s presentation. The child did not meet the case definition of MIS-C, as she did not have multiorgan dysfunction. Nevertheless, her positive IgM titer suggests acute SARS-CoV-2 infection could be an infectious trigger of KD in the absence of clinical findings of MIS-C.

Live-virus neutralization assays were performed using an mNeonGreen fluorescently labeled SARS-CoV-2 virus (Fig. 1C). Whereas all children with MIS-C had detectable neutralization titers to SARS-CoV-2, only 30% of children with COVID-19 had neutralizing antibodies. Children with MIS-C had significantly higher neutralizing antibody titers than children with COVID-19, KD, or hospitalized controls. RBD IgG binding titers strongly correlated with live-virus neutralization titers as determined by linear regression of the log-transformed titers (R^2^=0.667, P<0.001) (Fig. 1D).

We similarly performed linear regression analyses to identify correlations between RBD IgG antibody titers and MIS-C clinical and laboratory features. We found that RBD IgG antibody titers correlated strongly with the peak erythrocyte sedimentation rate (ESR) obtained prior to intravenous immune globulin (IVIG) administration (n=8, R^2^=0.512, P=0.046) (Fig. 1E). RBD IgG antibody titers also correlated with hospital length of stay (LOS) (R^2^=0.548, P=0.014) and ICU LOS (R^2^=0.590, P=0.010) (Fig. 1F), which were collinear outcome variables. Because all children with MIS-C in our cohort required ICU admission and inotropes or vasopressors for shock, the correlation between antibody titers and these patient outcomes could not be ascertained.

## DISCUSSION

We demonstrated that all children with MIS-C had high titers of SARS-CoV-2 RBD IgG antibodies which correlated with live-virus neutralization. Children with MIS-C also had significantly higher RBD antibody titers than children with COVID-19, KD, or hospitalized controls. These results suggest that quantitative SARS-CoV-2 RBD IgG serology may be helpful in establishing the diagnosis of MIS-C and distinguishing it from other syndromes with similar clinical appearances. Quantitative serology may also have prognostic value, as SARS-CoV-2 RBD IgG strongly correlated with metrics of systemic inflammation (ESR) and clinical outcomes (hospital and ICU LOS). The RBD antigen may be a well suited for quantitative antibody measurements due to its potent immunogenicity^14^ and lack of cross-reactivity with endemic coronavirus strains.^15^

Quantitative serology may also provide clues about MIS-C disease pathogenesis and about the timing of MIS-C following SARS-CoV-2 infection. To date, the pathogenesis of MIS- C is incompletely understood. Its temporal association following peak SARS-CoV-2 transmission in certain regions has led some to hypothesize MIS-C is attributable to post-infectious immune dysregulation and hyperinflammation.^16^ Interestingly, no children with MIS- C in our study recalled having an antecedent febrile or respiratory illness. However, all had detectable RBD IgM antibody titers, indicating a recent SARS-CoV-2 infection. Serologic studies in adults have demonstrated that SARS-CoV-2 antibody titers correlate with COVID-19 disease severity.^17-20^ Thus, it would seem paradoxical that children with asymptomatic infections would develop such robust immune responses as demonstrated by high titers of RBD IgG and neutralizing antibody responses. The reasons why some children develop MIS-C, while others do not is currently unknown. However, geographic, racial, and sociodemographic discrepancies suggest both genetic and environmental factors may play a role.

Our study has several limitations including a small sample size. Results should be corroborated in a larger cohort representing varying clinical phenotypes. Second, detection of SARS-CoV-2 binding and neutralizing antibodies in children with MIS-C demonstrates association, but not necessarily causation. Also, as the seroprevalence of SARS-CoV-2 antibodies increases, the clinical significance of these antibodies may eventually become obscured. Several patients with MIS-C had specimens drawn after administration of IVIG, which conceivably could have affected SARS-CoV-2 antibody titers. However, 4 of 5 patients with KD also received IVIG prior to specimen collection, with negligible effects on RBD titers. A recent study demonstrated various IVIG products lack cross-reactivity with the SARS-CoV-2 RBD.^15^

In conclusion, we found that all children with MIS-C had high titers of SARS-CoV-2 RBD IgG antibodies which correlated with neutralization. RBD IgG antibodies also correlated with metrics of systemic inflammation and with clinical outcomes. Thus, measuring quantitative SARS-CoV-2 RBD antibody titers may have a role in establishing the diagnosis of MIS-C, distinguishing it from other similar clinical entities, and stratifying risk for adverse outcomes.

## Data Availability

All data referred to in the manuscript can be made available upon request.

## ACKNOWLEDGEMENTS

The authors gratefully acknowledge our colleagues in clinical care, the clinical and research laboratories, and the clinical research nurses and coordinators, without whom this study would not have been possible. We thank all the study participants and their families, and we thank our families for their steadfast support.

## Abbreviations

AHA: American Heart Association
ANOVA: Analysis of variance
BMI: Body mass index
CDC: Centers for Disease Control and Prevention
COVID-19: Coronavirus disease 2019
ELISA: Enzyme-linked immunosorbent assay
ELISPOT: Enzyme-linked immune absorbent spot
ESR: Erythrocyte sedimentation rate
FRNT: Focus reduction neutralization titer
HFNC: High-flow nasal cannula
ICU: Intensive care unit
IVIG: Intravenous immune globulin
KD: Kawasaki Disease
LOD: Limit of detection
LOS: Length of stay
MIS-C: Multisystem inflammatory syndrome in children
N protein: Nucleocapsid protein
NP: Nasopharyngeal
RBD: Receptor binding domain
RT-PCR: Real-time polymerase chain reaction
S protein: Spike protein
SARS-CoV-2: Severe acute respiratory syndrome coronavirus 2

